# Epidemiological Characteristics of COVID-19; a Systemic Review and Meta-Analysis

**DOI:** 10.1101/2020.04.01.20050138

**Authors:** Malahat Khalili, Mohammad Karamouzian, Naser Nasiri, Sara Javadi, Ali Mirzazadeh, Hamid Sharifi

**Affiliations:** HIV/STI Surveillance Research Center, and WHO Collaborating Center for HIV Surveillance, Institute for Futures Studies in Health, Kerman University of Medical Sciences, Kerman, Iran; Department of Biostatistics and Epidemiology, School of Public Health, Kerman University of Medical Sciences, Kerman, Iran; School of Population and Public Health, University of British Columbia, Vancouver, BC, Canada; Modeling in Health Research Center, Institute for Futures Studies in Health, Kerman University of Medical Sciences, Kerman, Iran; Department of Epidemiology and Biostatistics, Institute for Global Health Sciences, University of California San Francisco, San Francisco, CA, USA

## Abstract

**Background:** Our understanding of the corona virus disease 2019 (COVID-19) continues to evolve. However, there are many unknowns about its epidemiology.

**Purpose:** To synthesize the number of deaths from confirmed COVID-19 cases, incubation period, as well as time from onset of COVID-19 symptoms to first medical visit, ICU admission, recovery and death of COVID-19.

**Data Sources:** MEDLINE, Embase, and Google Scholar from December 01, 2019 through to March 11, 2020 without language restrictions as well as bibliographies of relevant articles.

**Study Selection:** Quantitative studies that recruited people living with or died due to COVID-19.

**Data Extraction:** Two independent reviewers extracted the data. Conflicts were resolved through discussion with a senior author.

**Data Synthesis:** Out of 1675 non-duplicate studies identified, 57 were included. Pooled mean incubation period was 5.84 (99% CI: 4.83, 6.85) days. Pooled mean number of days from the onset of COVID-19 symptoms to first clinical visit was 4.82 (95% CI: 3.48, 6.15), ICU admission was 10.48 (95% CI: 9.80, 11.16), recovery was 17.76 (95% CI: 12.64, 22.87), and until death was 15.93 (95% CI: 13.07, 18.79). Pooled probability of COVID-19-related death was 0.02 (95% CI: 0.02, 0.03).

**Limitations:** Studies are observational and findings are mainly based on studies that recruited patient from clinics and hospitals and so may be biased toward more severe cases.

**Conclusion:** We found that the incubation period and lag between the onset of symptoms and diagnosis of COVID-19 is longer than other respiratory viral infections including MERS and SARS; however, the current policy of 14 days of mandatory quarantine for everyone might be too conservative. Longer quarantine periods might be more justified for extreme cases.

**Funding:** None.

**Protocol registration:** Open Science Framework: https://osf.io/a3k94/

## Background

The severe acute respiratory syndrome coronavirus 2 (SARS-CoV-2) was first identified in a few unusual pneumonia patients linked to the Wuhan seafood wholesale market in China in December 2019 (1). However, it soon grew out of China and the corona virus disease 2019 (COVID-19) was declared a pandemic on March 11, 2020 and is now found in 190 countries (2). While the epidemic seems to have slowed down in China due to the strict quarantine and preventative regulations, the number of COVI-19 patients (i.e., >370,000 as of March 23^rd^) and confirmed deaths (i.e., i.e., >16,000 as of March 23^rd^) are rapidly increasing (2); figures that greatly surpass that of other diseases in the family of coronaviruses with similar genomes to SARS-CoV-2 such as severe acute respiratory syndrome (SARS) and Middle East Respiratory Syndrome (MERS) (3) which emerged in 2003, 8,098 patients and 774 deaths across 29 countries, and 2012, leading to 2494 patients and 858 deaths across 27 countries, respectively (4-6). The healthcare systems in many countries, such as Italy, Iran, Spain, and France are overwhelmed and struggling with the soaring number of patients (7).

Although our understanding of COVID-19’s epidemiology is evolving, it is assumed that SARS-CoV-2 is mainly transmitted via droplets and close contacts with people carrying the virus (2). However, recent reports have also proposed the possibility of the virus being contracted via various surfaces, gastrointestinal transmission (8), and potentially airborne (2, 9). Based on the existing evidence, elderly population, those with suppressed immune systems, and underlying metabolic, cardiovascular or respiratory diseases are at an increased risk for adverse outcomes; however, recent reports from outside China point to a considerable risk of severe outcomes among the general adult population (i.e., <65 years old) (10, 11).

As we continue to learn more about COVID-19 and its characteristics, there are many unknowns and confusions about its epidemiology such as hospitalization- and recovery-related outcomes which are indeed of utmost importance when it comes to health system preparedness (12, 13). For example, mean number of days of incubation for COVID-19 varies greatly across the existing literature ranging from 2.5 (14) to 12.1 (15) days. Our knowledge of time from contracting the disease to recovery or death are even more limited. In this systematic review, we tried to identify the studies that recruited patients diagnosed with COVID-19 and calculate polled estimates for several epidemiological and clinical outcomes to help provide an overall picture of the characteristics of COVID-19. Findings of this study could help inform the ongoing public health and public policy practices across the world.

## Methods

The details of inclusion criteria and analytical approach were designed a priori and are documented in Open Science Framework (https://osf.io/a3k94/).

### Databases and Search Strategy

Following the Systematic Reviews and Meta-Analyses (PRISMA) checklist (16) and the Peer Review of Electronic Search Strategies (PRESS) guideline (17), we searched PubMed, Embase, and Google Scholar from December 01, 2019 through to March 11, 2020 for studies that measured and reported several characteristics of COVID-19 (e.g., incubation period, hospitalization, death). Search terms were combined using appropriate Boolean operators and included subject heading terms/keywords relevant to COVID-19 (e.g., novel coronavirus, sars-cov2, coronavirus disease). Please see **supplement 1** for a sample search strategy.

### Inclusion criteria

Quantitative studies were included in the review if they reported incubation period or time from onset of the symptoms to first medical visit, intensive care unit (ICU) admission, recovery or death. Studies were also included if they reported number of deaths from patients with confirmed COVID-19 diagnosis. Studies were not excluded based on language, location, or measurement method. Given the nature of the study which used secondary data involving no interaction with humans, no ethics approval was required.

### Study Selection

Two authors (SJ and NN) completed the abstract and full-text screening, independently. Citations that met our eligibility criteria or were unclear were screened at full-text by two independent reviewers (SJ and NN) in duplicate. Disagreements over inclusion of studies were resolved through discussion. Duplicate records were excluded.

### Data Extraction

Data were extracted independently by the two authors (SJ and NN), and discrepancies were resolved through discussion or by arbitration with a senior co-author (AM). Data were extracted on study type (e.g., descriptive, case-series, mathematical modeling), publication year, location, sample size, patients’ age, gender, exposure history, X-ray and computed tomography (18) scan findings, symptoms, and underlying conditions if reported. We also extracted data on the main outcomes of interests including the number of deaths from confirmed COVID-19 cases, incubation period, as well as time from onset of COVID-19 symptoms to first medical visit, to ICU admission, to recovery and to death.

### Statistical analysis

For studies that did not provided enough data to be included in the meta-analysis, we performed a qualitative data synthesis. Case-reports with a sample size of one were also removed from the meta-analysis as they did not provide any dispersion estimate. Meta-analysis was performed using STATA’s (V.15.1) metan (for numerical variables) and metaprop (for binary variables) commands. The 95% confidence intervals (CI) for binary variables were computed using the exact binomial method. Heterogeneity between studies was assessed using both the I^2^ statistic with a cutoff of ≥ 50% and the Chi-square test with P-value <0.10 to define a significant degree of heterogeneity (19). As all results turned to be significantly heterogenous, we used random-effects model to calculate the pooled point estimate and 95% CI for the mortality rate, mean time from onset of COVID-19 symptoms to first medical visit, ICU admission, recovery and death. For mean incubation period, we estimated 99% CI. We also conducted a random-effects meta-regression using STATA’s metareg command to identify the sources of heterogeneity and explore the effect of study-level covariates where data were available. A two-sided P-value <0.05 was considered as statistically significant effect.

## Results

### Participants and study characteristics

We found a total of 1675 non-duplicate studies, 57 of which were included in our qualitative synthesis and 43 were considered for meta-analysis (Figure 1). A description of the main characteristics of the included studies is provided in Table 1. The 57 studies included 27 cross-sectional, one case-control, one retrospective cohort, and 28 case series/case report studies with sample sizes of observational studies ranging greatly from one to 58182 for a study in the Hubei province (20). Inclusion criteria varied greatly across the studies but most participants were hospitalized patients living or traveling from various provinces in China. Median (range) age of the participants was 46.2 (17 days to 78.5 years) and about 60% were men. Most studies were conducted between January and February 2020. Clinical and epidemiological characteristics of the patients included in the study are presented in Table 2. Among studies that reported exposure history among their participants, most patients were directly or indirectly traced back to the city Wuhan (e.g., lived in Wuhan or had recently travelled to Wuhan) and the Huanan seafood market in Hubei province, China. Several cases of contracting COVID-19 through close contacts with family members were also reported across the studies. Frequent CT or X-ray findings included thickened texture of lungs, bilateral focal consolidation, lobar consolidation, ground glass opacity, and patchy consolidation, and unilateral/bilateral pneumonia. Common symptoms reported across the studies include fever, cough, shortness of breath, and fatigue/weakness. Only 15 studies reported some information about the pre-existing conditions of the patients; most of whom had metabolic and cardiovascular underlying conditions.

**Table 1.** Characteristics of the included studies in the systematic review

| First Author | Publication Date<br>(DD-MM-YY) | Study Setting/Location | Study Type | Sample size | Age; Mean/Range | Male Proportion |
| --- | --- | --- | --- | --- | --- | --- |
| Chan (39) | 24-Jan-20 | China, Shenzhen | Case series | 6 | 46.17 | 0.50 |
| Li (24) | 29-Jan-20 | China, Wuhan | Cross-sectional | 425 | 55.5 | 0.56 |
| Chen (1) | 30-Jan-20 | China, Wuhan | Cross-sectional | 99 | 55.5 | 0.68 |
| Holshue (40) | 31-Jan-20 | USA, Washington | Case-report<sup>a</sup> | 1 | 35 | 1.00 |
| Wu (41) | 3-Feb-20 | China, Wuhan | Case-report | 1 | 41 | 1.00 |
| Kim (42) | 3-Feb-20 | Korea, Incheon | Case-report | 1 | 35 | 0.00 |
| Cai (43) | 4-Feb-20 | China, Shanghai | Case-report<sup>b</sup> | 2 | 7 | 0.67 |
| Lin (44) | 4-Feb-20 | China, Jiangxi province | Case-report | 2 | 37 | 1.00 |
| Backer (26) | 6-Feb-20 | China, Wuhan | Cross-sectional | 88 | 2 to 72 | 0.65 |
| Ki (45) | 9-Feb-20 | South Korea | Cross-sectional | 28 | 42.1 | 0.54 |
| Jiang (46) | 24-Feb-20 | China | Cross-sectional | 50 | NR | NR |
| Chen (47) | 11-Feb-20 | China, Wuhan | Case-report | 1 | 1 | 1.00 |
| Thompson (48) | 11-Feb-20 | China | Cross-sectional | 47 | 47 | 0.63 |
| Zhang (49) | 11-Feb-20 | China, Xiaogan | Case-report | 1 | 0.25<br>(3 months) | 0.00 |
| Duan (50) | 12-Feb-20 | China, Wuhan | Case-report | 1 | 46 | 0.00 |
| Stoecklin (51) | 13-Feb-20 | France, Bordeaux | Case-report | 3 | 36.3 | 0.67 |
| Huang (52) | 15-Feb-20 | China, Wuhan | Cross-sectional | 41 | 49.34 | 0.73 |
| Zhang (14) | 15-Feb-20 | China, Beijing | Case series | 9 | 35.3 | 0.56 |
| Lim (53) | 17-Feb-20 | Korea, Goyang | Case-report | 1 | 54 | 1.00 |
| Linton (54) | 17-Feb-20 | China, multiple cities | Cross-sectional | 276 | 30–59 (>50%) | 0.58 |
| COVID-19 Response Team (36) | 17-Feb-20 | China, multiple cities | Cross-sectional | 44672 | 30-69 (77.8%) | 0.51 |
| Zeng (55) | 17-Feb-20 | China, Wuhan | Case-report | 1 | 0.046<br>(17 days) | 1.00 |
| Yu (56) | 18-Feb-20 | China, Shanghai | Case-report | 4 | 74.25 | 0.50 |
| Xu (57) | 18-Feb-20 | China, Beijing | Case-report | 1 | 50 | 1.00 |
| Fang (58) | 19-Feb-20 | China, Chengdu | Case-report | 1 | 47 | 1.00 |
| Wang (59) | 20-Feb-20 | China, Wuhan | Cross-sectional | 138 | 55.3 | 0.54 |
| Zhu (60) | 20-Feb-20 | China, Wuhan | Case-report | 3 | 47.33 | 0.67 |
| Bai (15) | 21-Feb-20 | China, Anyang | Case series | 6 | 34.75 | 0.17 |
| Hao (61) | 21-Feb-20 | China, Shaanxi | Case-report | 1 | 58 | 1.00 |
| Shi (62) | 24-Feb-20 | China, Wuhan | Cross-sectional | 81 | 49.5 | 0.52 |
| **Cheng (63)** | 26-Feb-20 | Taiwan, Taoyuan | Case-report | 1 | 55 | 0.00 |
| **Li (64)** | 26-Feb-20 | China, Mainland China | Cross-sectional | 44653 | NR | NR |
|  | 26-Feb-20 | China, Hubei | Cross-sectional | 33366 | NR | NR |
| **Yang (65)** | 26-Feb-20 | China, Wenzhou | Cohort | 149 | 45.11 | 0.54 |
| **Shrestha (66)** | 27-Feb-20 | Nepal, Kathmandu | Case-report | 1 | 32 | 1.00 |
| **Tian (67)** | 27-Feb-20 | China, Beijing | Cross-sectional | 262 | 47.5 | 0.48 |
| **Guan (68)** | 28-Feb-20 | China, 30 provinces | Cross-sectional | 1099 | 46.7 | 0.58 |
| **Lillie (69)** | 28-Feb-20 | UK | Case-report | 2 | 36.5 | 0.50 |
| **Wu (70)** | 29-Feb-20 | China, Jiangsu province | Cross-sectional | 80 | 46.1 | 0.49 |
| **Song (71)** | 1-Mar-20 | China | Cross-sectional | 11791 | NR | NR |
| **Cheng (72)** | 2-Mar-20 | China, Henan province | Cross-sectional | 1079 | 46.6 | 0.53 |
| **Peng (73)** | 2-Mar-20 | China, Wuhan | Cross-sectional | 112 | 61.3 | 0.47 |
| **Dey (20)** | 3-Mar-20 | China, Hubei province | Cross-sectional | 58182 | NR | NR |
|  |  | China, Other provinces | Cross-sectional | 12264 | NR | NR |
|  |  | Outside of China | Cross-sectional | 425 | NR | NR |
| **Ruan (74)** | 3-Mar-20 | China, Wuhan | Case control | 66 | 45-75 (>50%) | NR |
| **Young (75)** | 3-Mar-20 | Singapore | Cross-sectional | 18 | 49.5 | 0.50 |
| **Chen (76)** | 5-Mar-20 | China, Guangdong | Case-report | 1 | 46 | 0.00 |
| **Cheng (77)** | 5-Mar-20 | China, Hong Kong | Cross-sectional | 42 | 57.8 | 0.48 |
| **Qiu (78)** | 5-Mar-20 | China, Zhengzhou | Case series | 8 | 25.9 | 0.50 |
| **Rothe (79)** | 5-Mar-20 | Germany, Munich | Case series | 5 | NR | NR |
| **Spiteri (80)** | 5-Mar-20 | WHO European Region | Cross-sectional | 38 | 41.75 | 0.66 |
| **Wang (81)** | 5-Mar-20 | China, Wuhan | Case-report | 2 | 78.5 | 0.50 |
| **Wang (82)** | 5-Mar-20 | China, Zhengzhou | Cross-sectional | 18 | 41 | 0.56 |
| **Wu (83)** | 5-Mar-20 | China, Tianjin | Cross-sectional | 40 | 44.0 | 0.33 |
| **Yang (84)** | 5-Mar-20 | China | Cross-sectional | 325 | 8 months to 90 years | 0.49 |
| **Lauer (27)** | 10-Mar-20 | China, outside Hubei province | Cross-sectional | 181 | 44.67 | 0.60 |
|  |  | Outside mainland China | Cross-sectional | 108 | NR | NR |
| **Tong (85)** | 17-May-20<sup>c</sup> | China, Zhoushan | Case-report | 3 | 33 | 1.00 |
| **Arashiro (86)** | 17-Jun-20<sup>c</sup> | Japan, Cruise ship | Case-report | 2 | 31 | 0.50 |
| **Liu (87)** | 17-Jun-20<sup>c</sup> | China, Shenzhen | Cross-sectional | 365 | 46.2 | 0.50 |

<sup>a</sup> Studies with a sample size less than or equal to 4 patients were labeled as case-reports (88); <sup>b</sup> A 7-year-old-boy and his parents; <sup>c</sup> Studies are in press and will be published in future issues of the respective journal

**Table 2.**
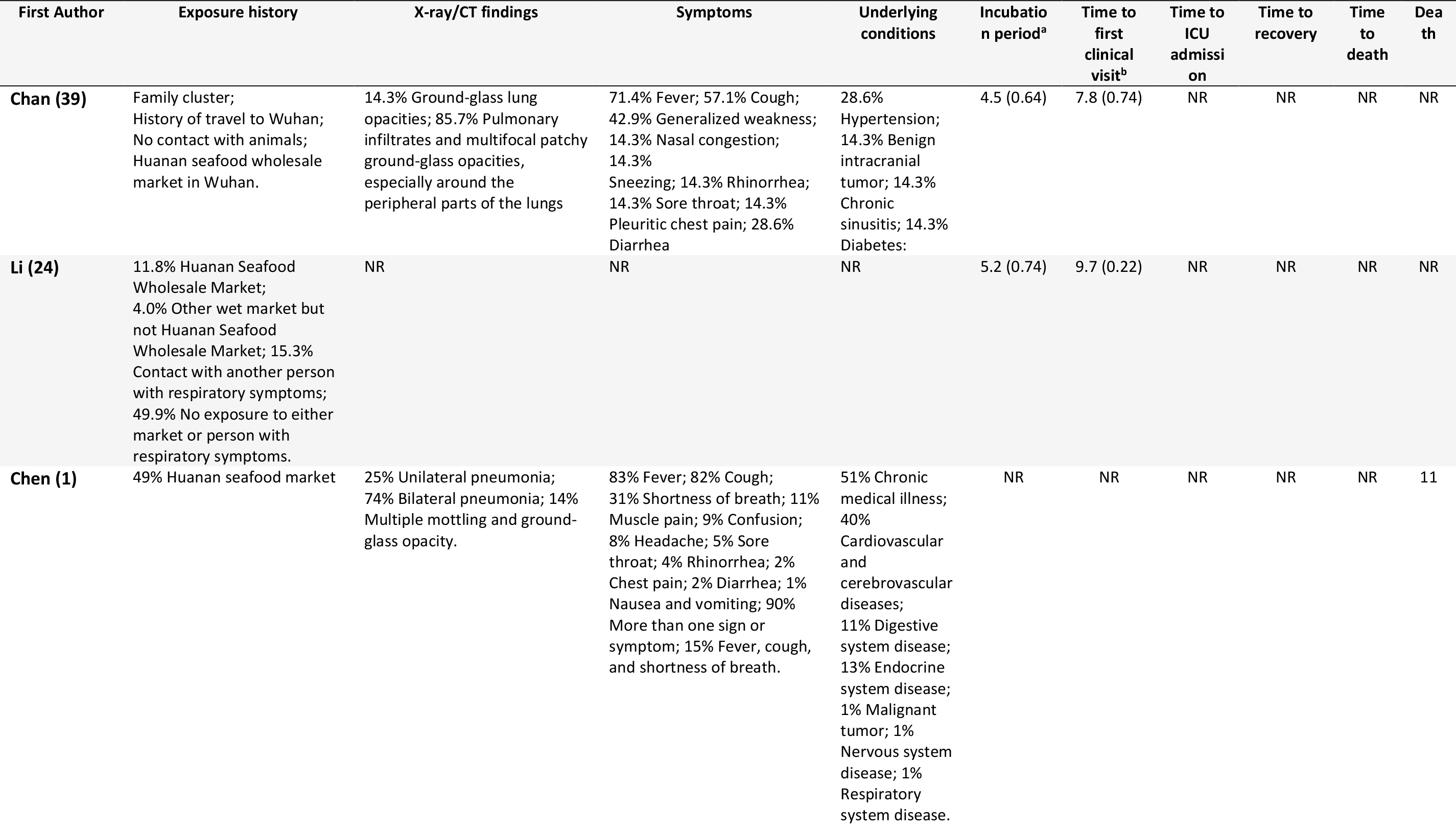
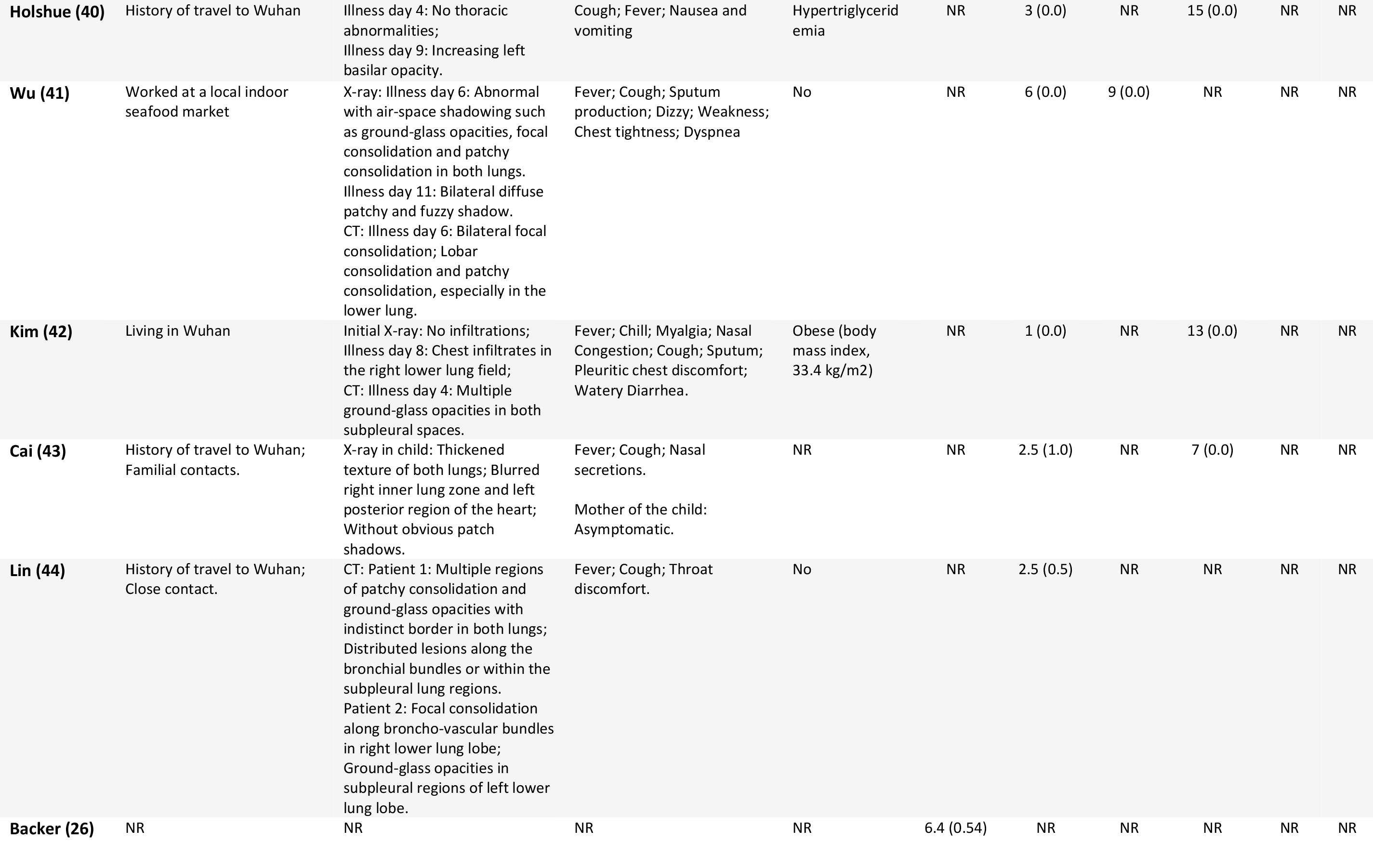
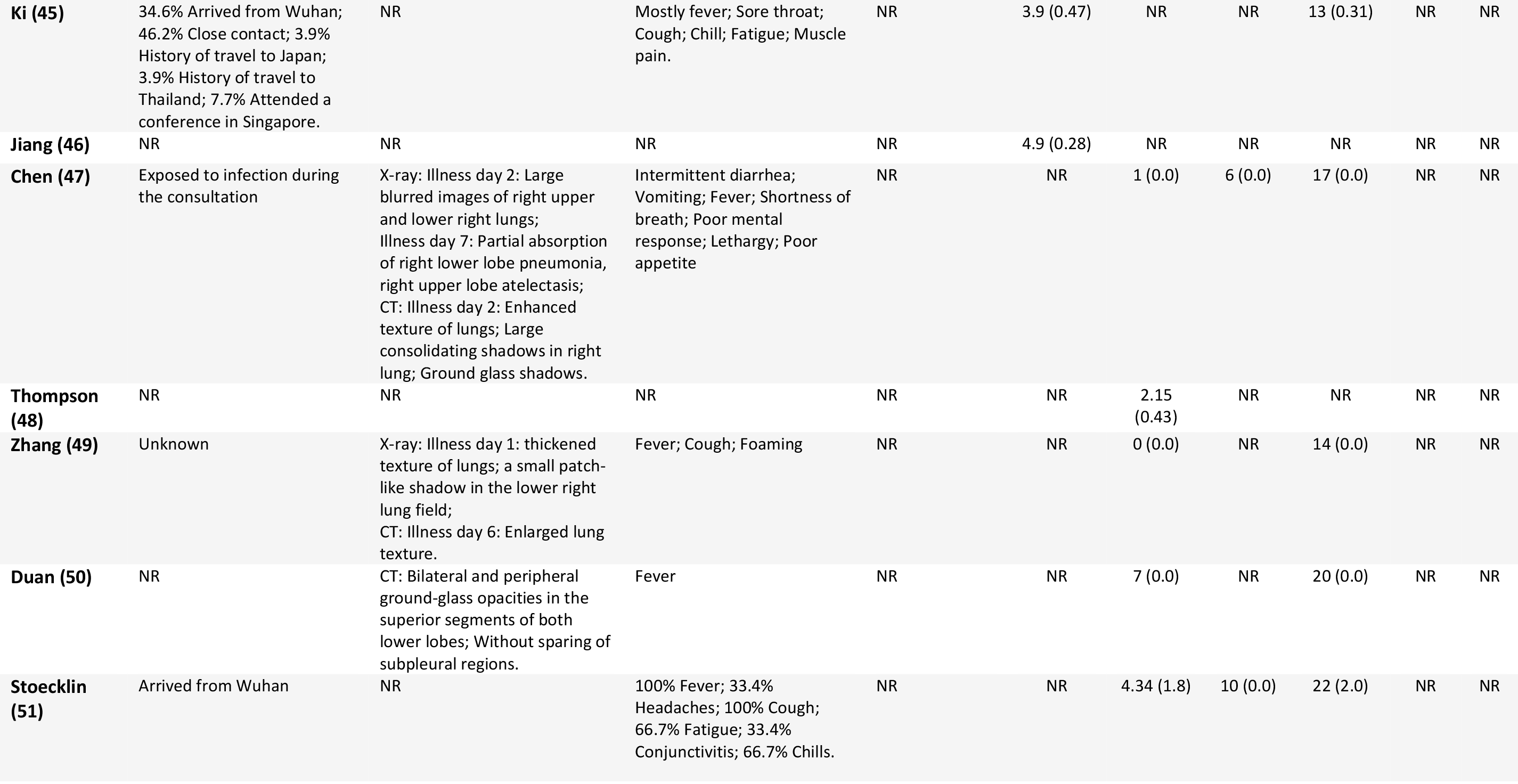
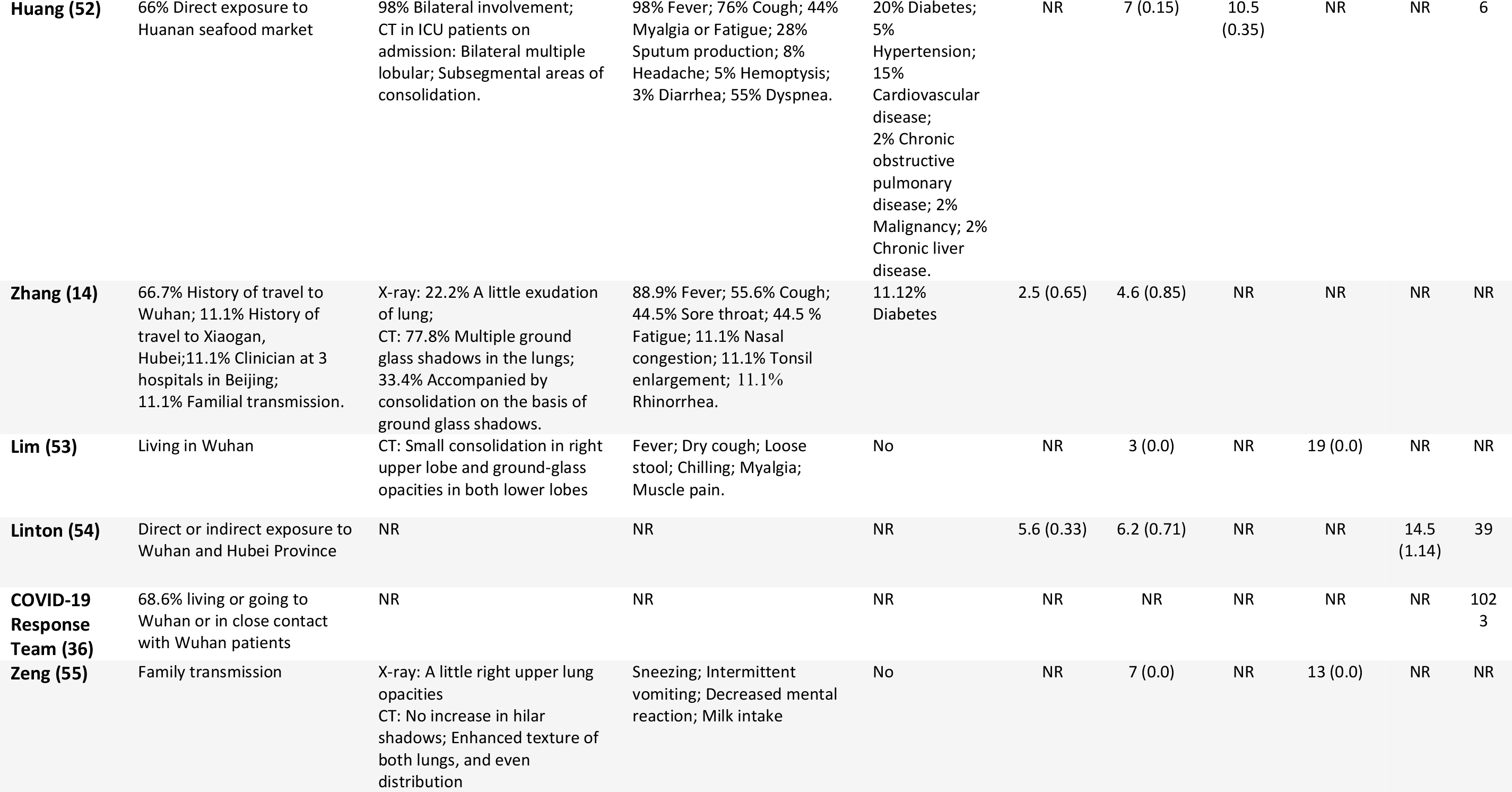
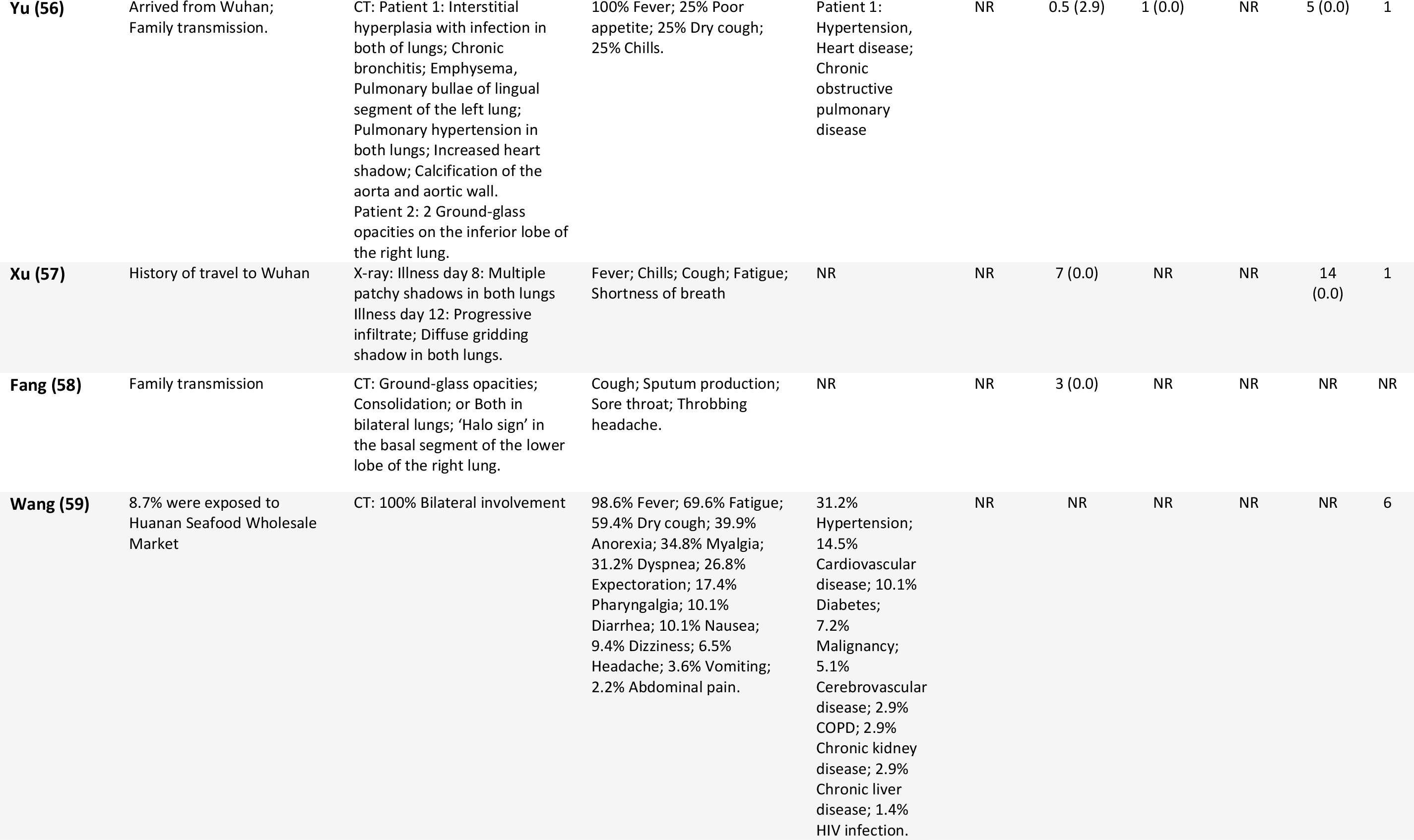
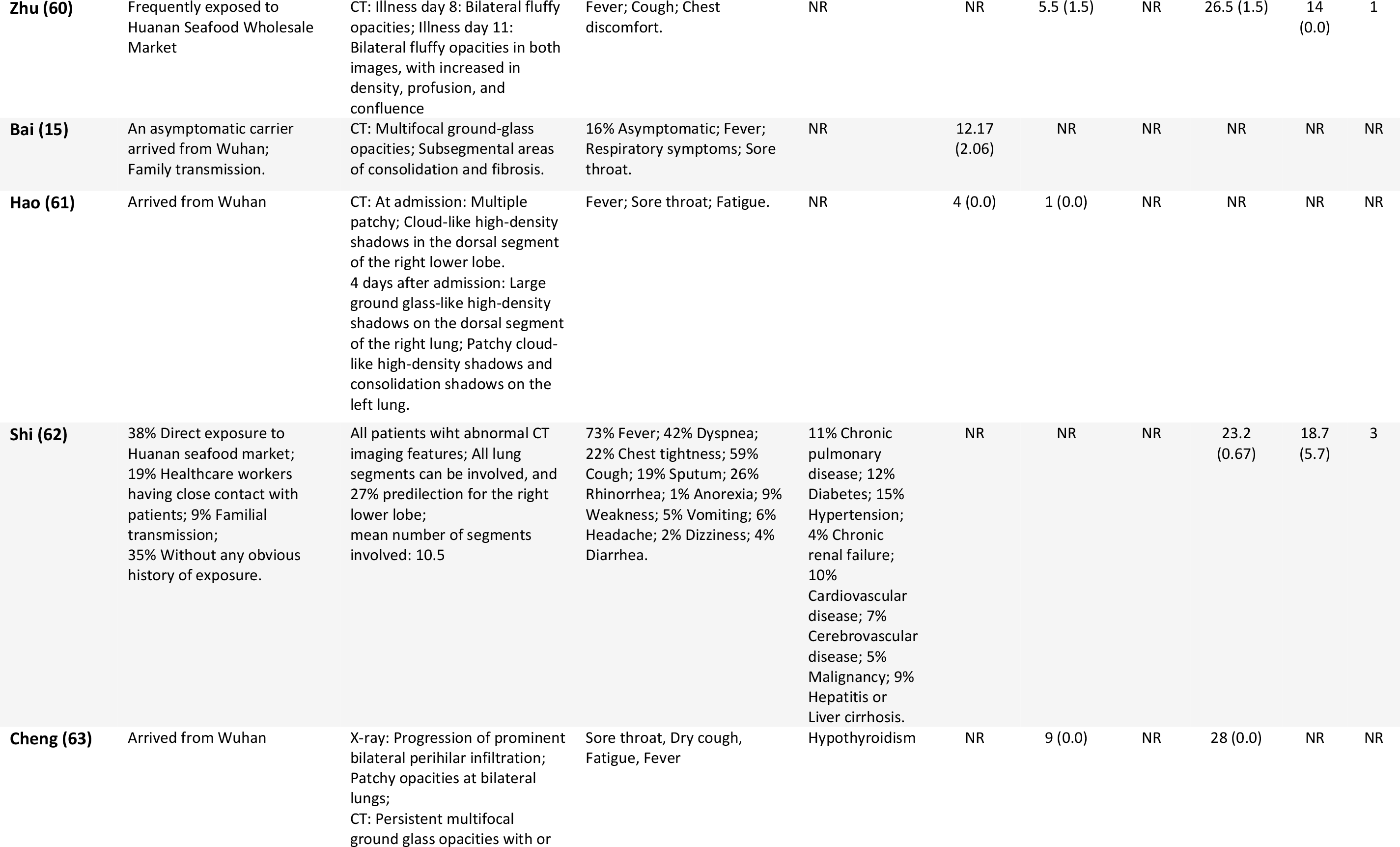
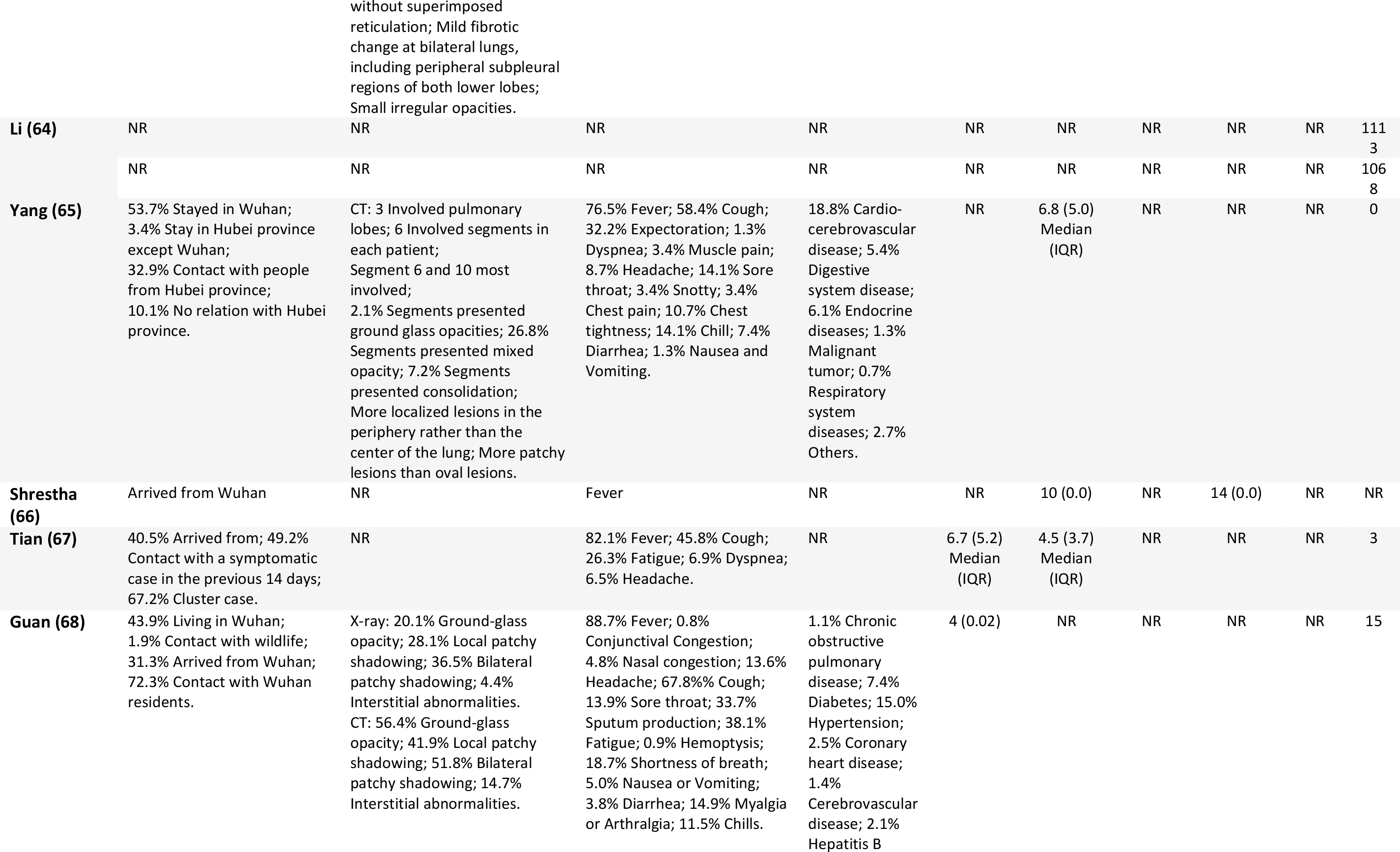
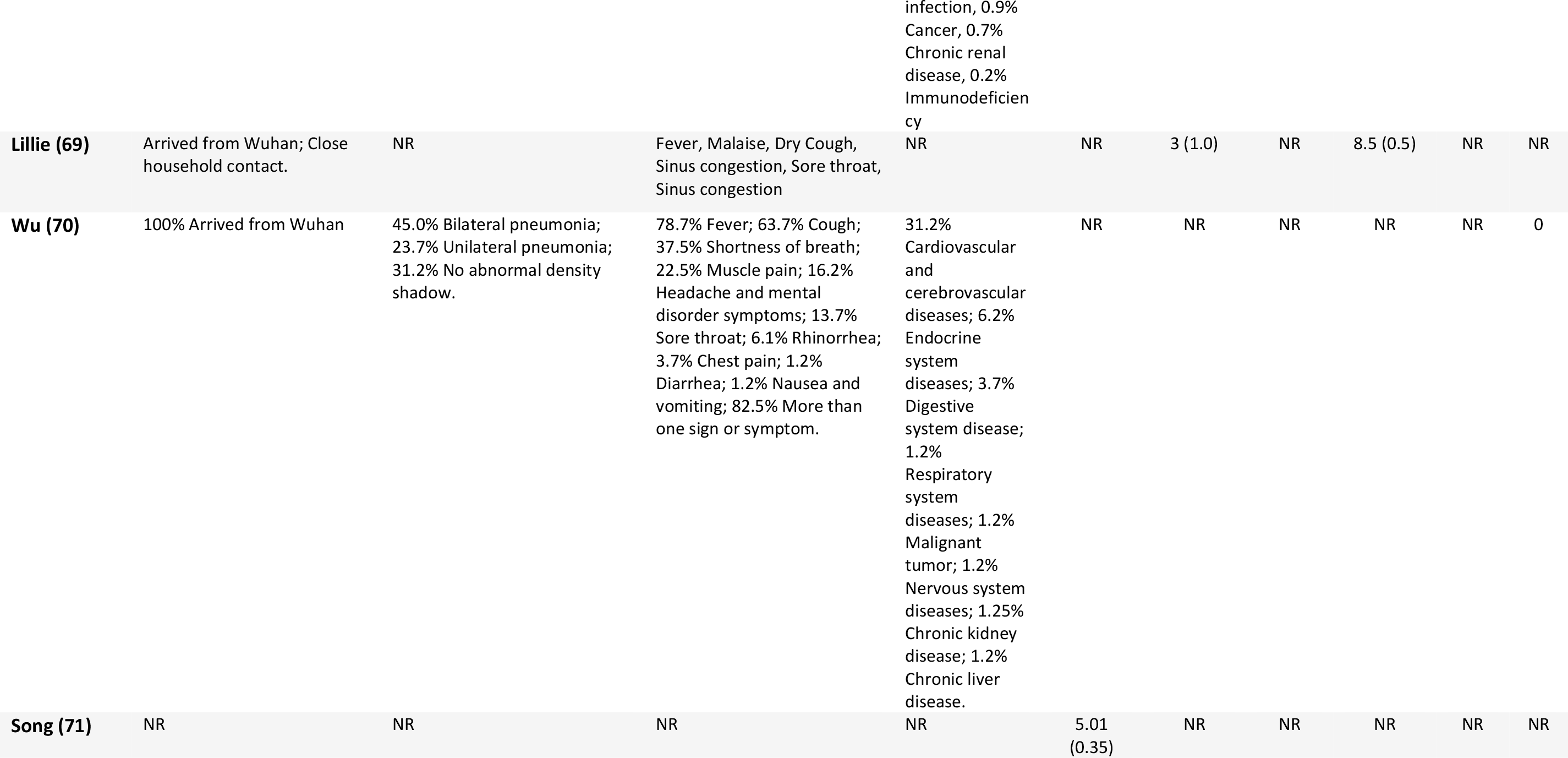
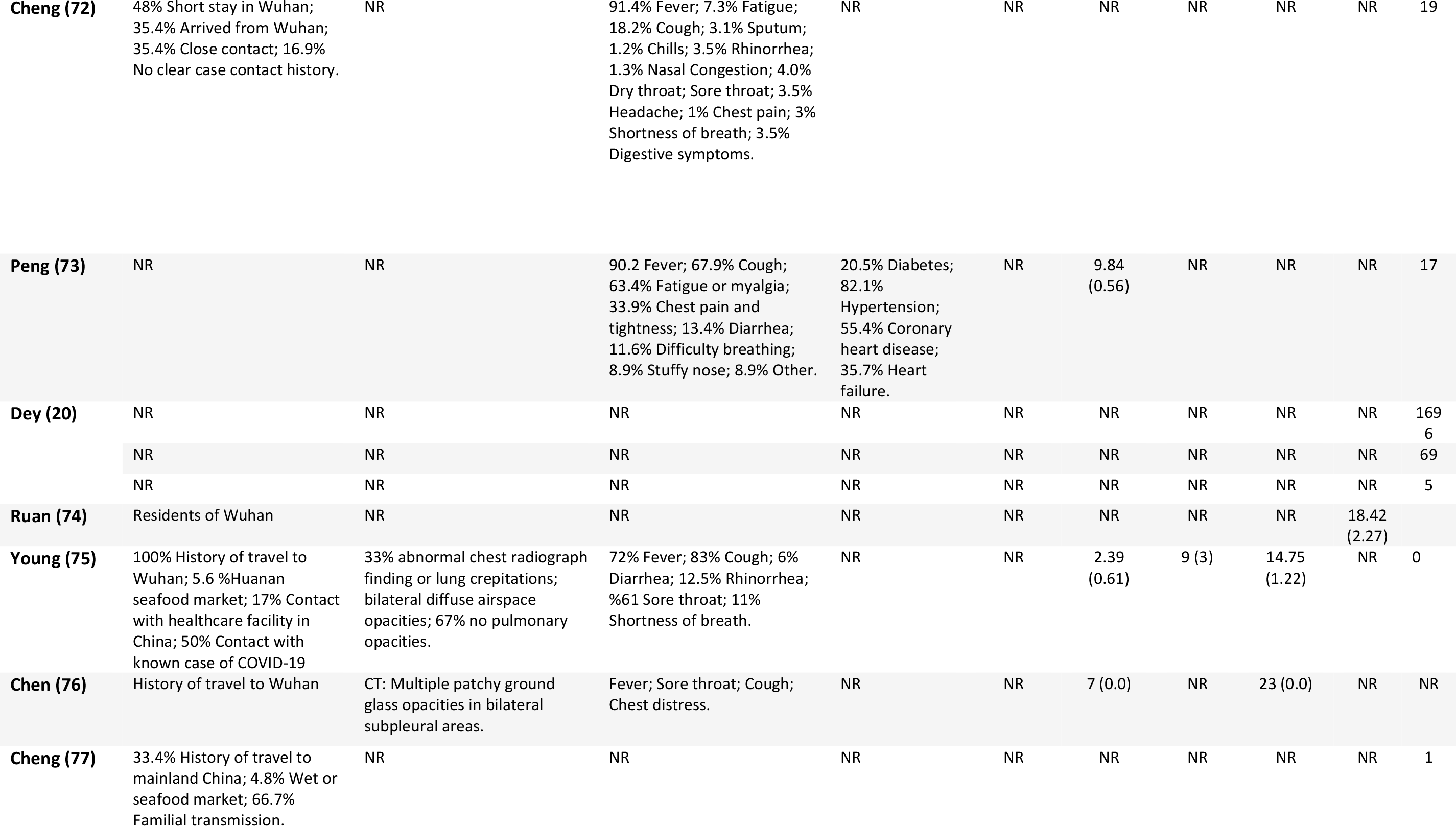
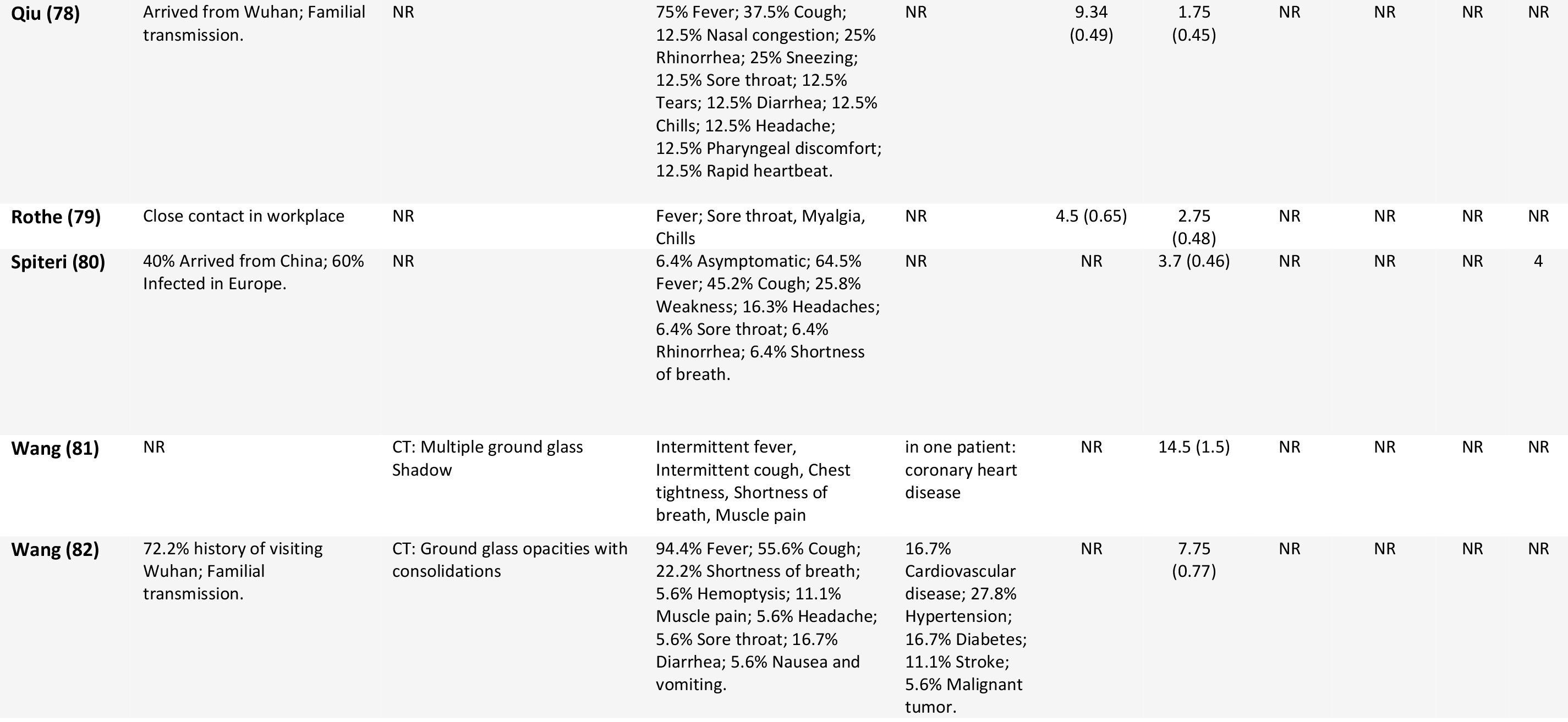
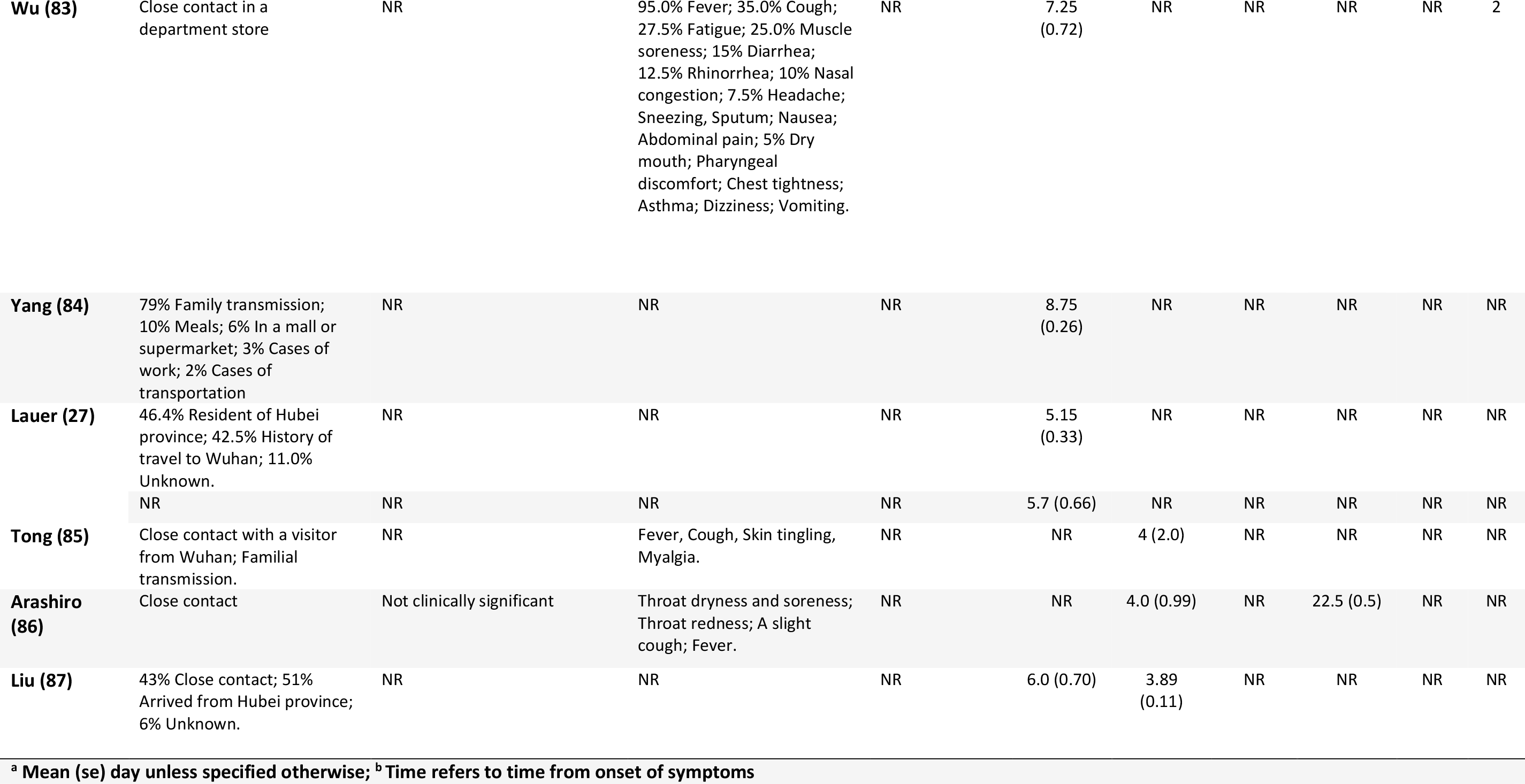
Medical and epidemiological characteristics of studies included in the review

**Figure 1.**
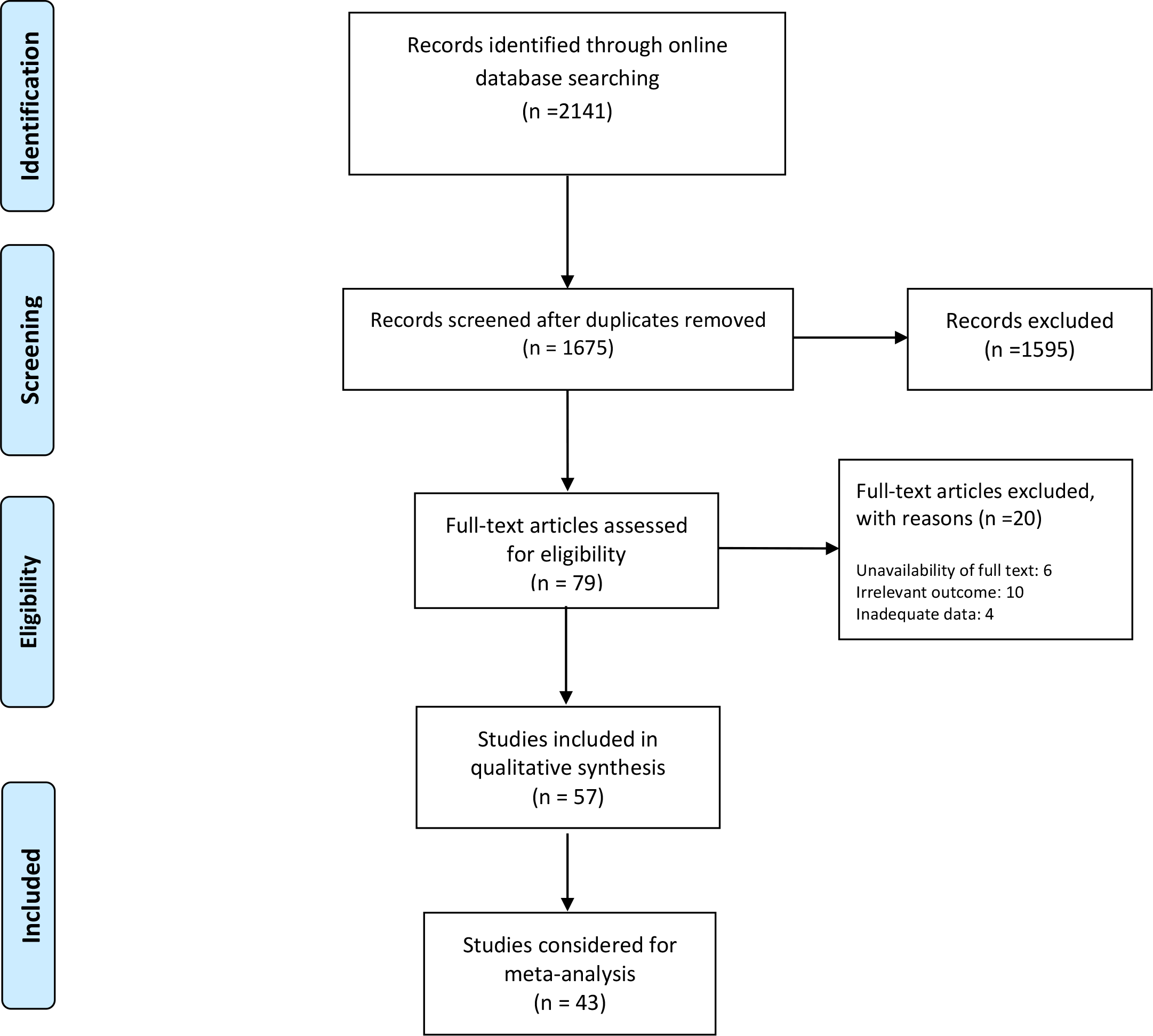
PRISMA flowchart of screened and included studies

### Mean incubation period

The estimated mean incubation period obtained from the included studies as well as resulting pooled mean are presented in Fig. 2. Out of the 14 studies included in the meta-analysis, 12 were conducted in China. The findings of meta-analysis showed that the mean incubation period was 5.84 (95% CI: 4.83, 6.85) days. Heterogeneity testing (I^2^=94.7%) revealed notable differences among the included studies in the meta-analysis. Multivariate meta-regression results (**Table 2**) showed no significant differences in incubation period time by country (China vs. others, Adjusted Beta = 1.70; P-value = 0.484), age (Adjusted Beta = -1.16; P-value = 0.239) or percent of male (Adjusted Beta = -13.07; P-value = 0.09) participants.

**Figure 2.**
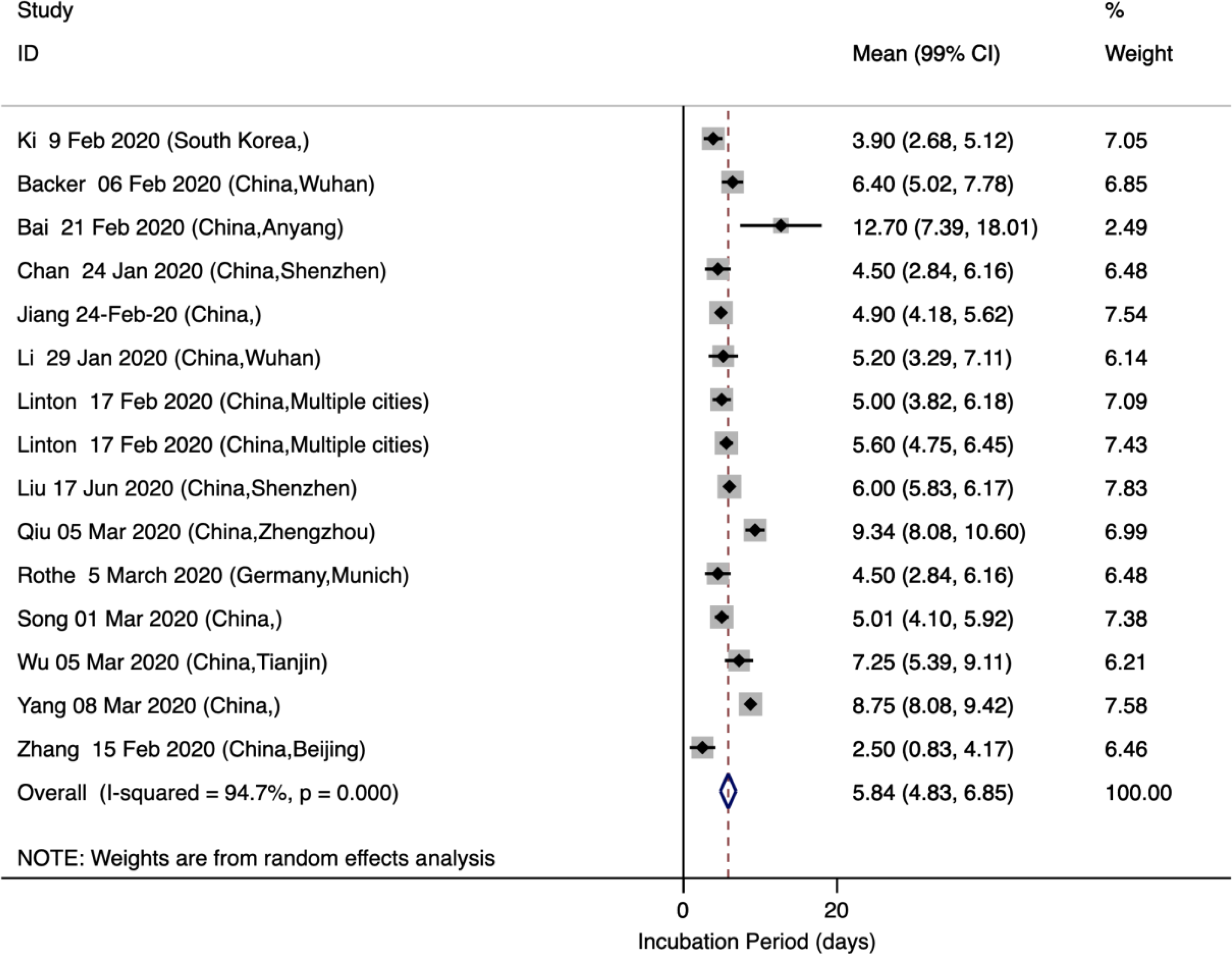
Incubation period of COVID-19 infection

### Mean time from onset of symptoms to first clinical visit

The estimated mean number of days from the onset of COVID-19 symptoms to first clinical visit was 4.82 (95% CI: 3.48, 6.15). As shown in Fig. 3, out of the 21 studies included in the meta-analysis, only 6 were conducted outside China. Heterogeneity testing (I^2^=98.0%) revealed notable differences among the included studies in the meta-analysis. Multivariate meta-regression results (**Supplement 2**) showed no significant differences in time from onset of symptoms to first clinical visit by country (China vs. others, Adjusted Beta = 1.47; P-value = 0.456), age (Adjusted Beta = 0.91; P-value = 0.187) or percent of male (Adjusted Beta = -2.96; P-value = 0.614) participants.

**Figure 3.**
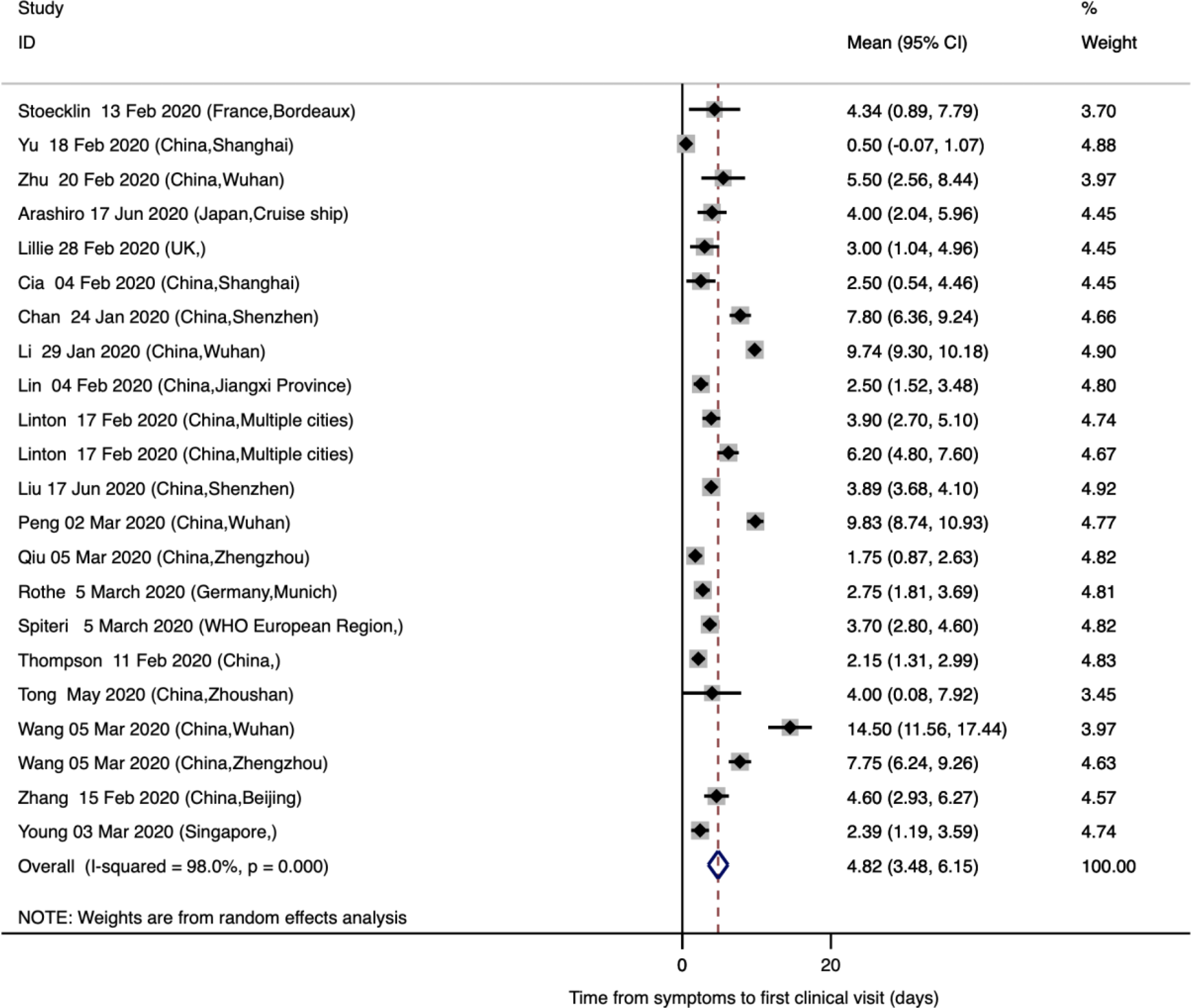
Time from onset of symptoms to first clinical visit for COVID-19 infection

### Mean time from onset of symptoms to ICU admission

The estimated mean number of days from the onset of COVID-19 symptoms to ICU admission was 10.48 (95% CI: 9.80, 11.16), an estimate that was derived from one study in Singapore and one in Wuhan, China (Fig. 4).

**Figure 4.**
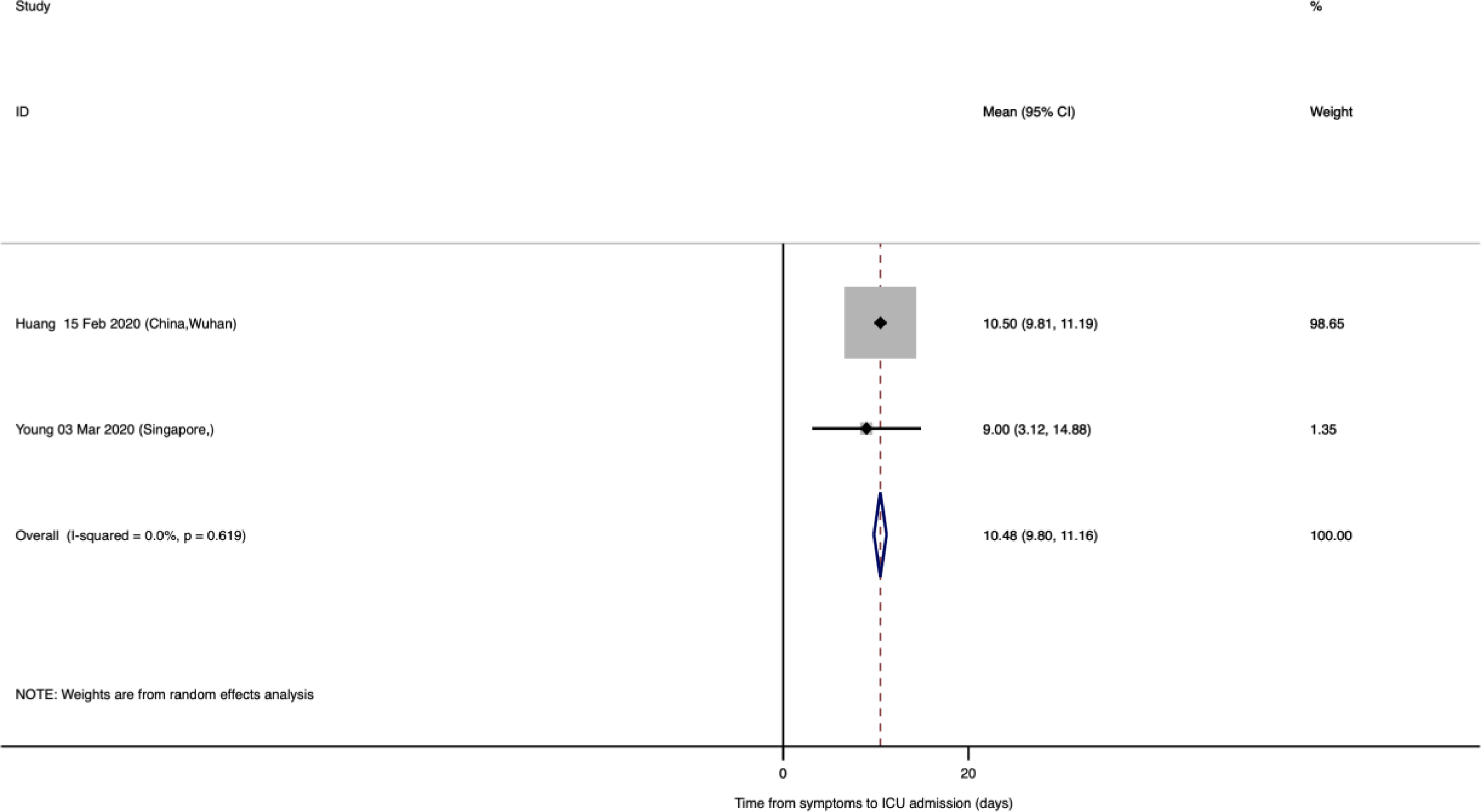
Time from onset of symptoms to ICU admission for COVID-19 infection

### Mean time from onset of symptoms to recovery

The estimated mean number of days from the onset of symptoms to recovery was reported in 6 studies and the resulting pooled mean was 17.76 (95% CI: 12.64, 22.87). Only one of the studies was conducted in China and the rest were completed in France, South Korea, UK, and Japan (Fig. 5). Heterogeneity testing (I^2^=99.0%) revealed notable differences among the included studies in the meta-analysis. Multivariate meta-regression results (**Supplement 2**) showed no significant differences in time from onset of symptoms to recovery by country (China vs. others, Adjusted Beta = 8.40; P-value = 0.522), age (Adjusted Beta = -3.02; P-value = 0.602) or percent of male (Adjusted Beta = 35.56; P-value = 0.537) participants.

**Figure 5.**
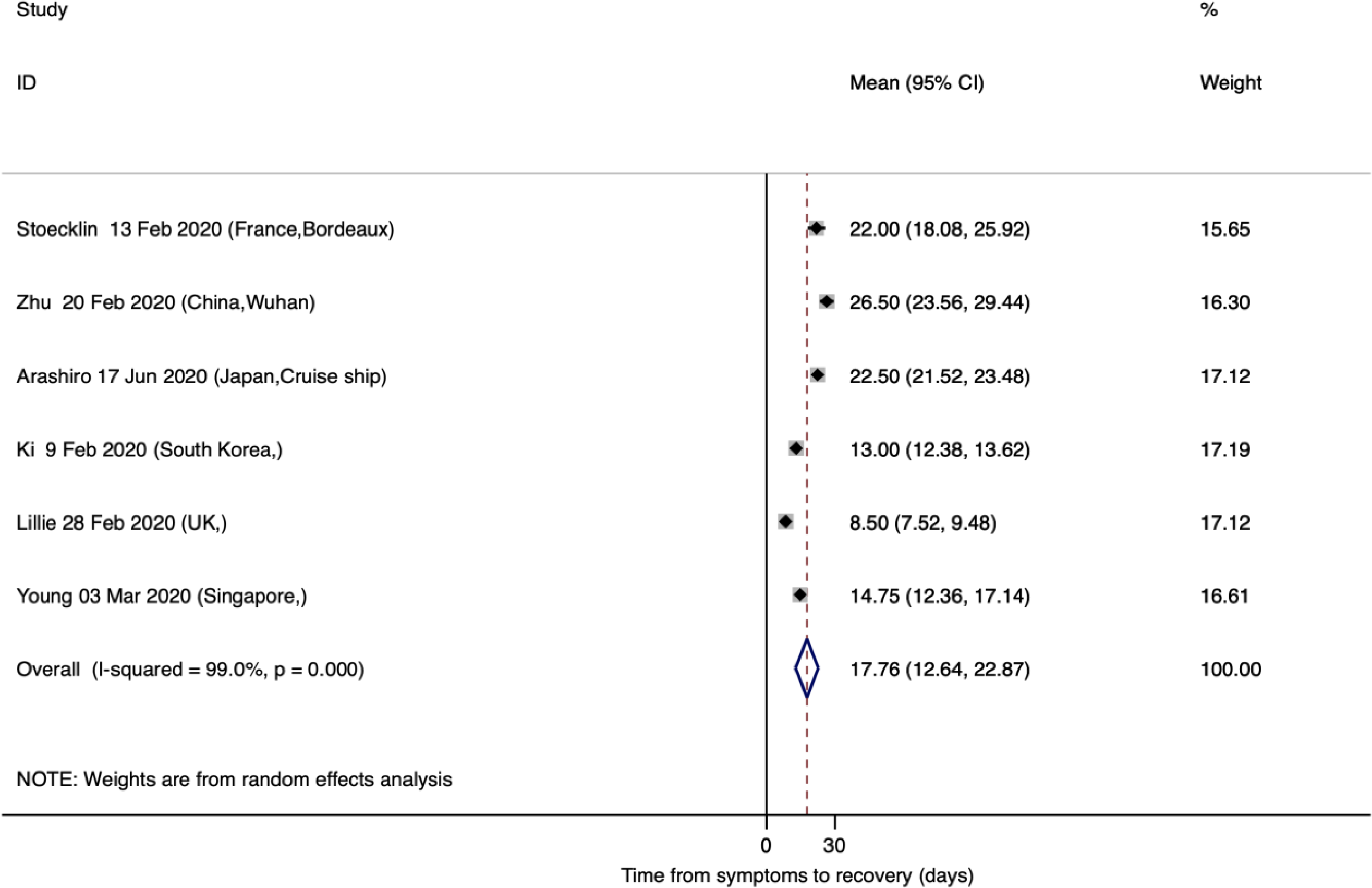
Time from onset of symptoms to recovery for COVID-19 infection

### Mean time from onset of symptoms to death

The estimated mean number of days from the onset of symptoms to death was reported in three studies with a pooled mean of 15.93 (95% CI: 13.07, 18.79). All of the studies were conducted in China (Fig. 6).

**Figure 6.**
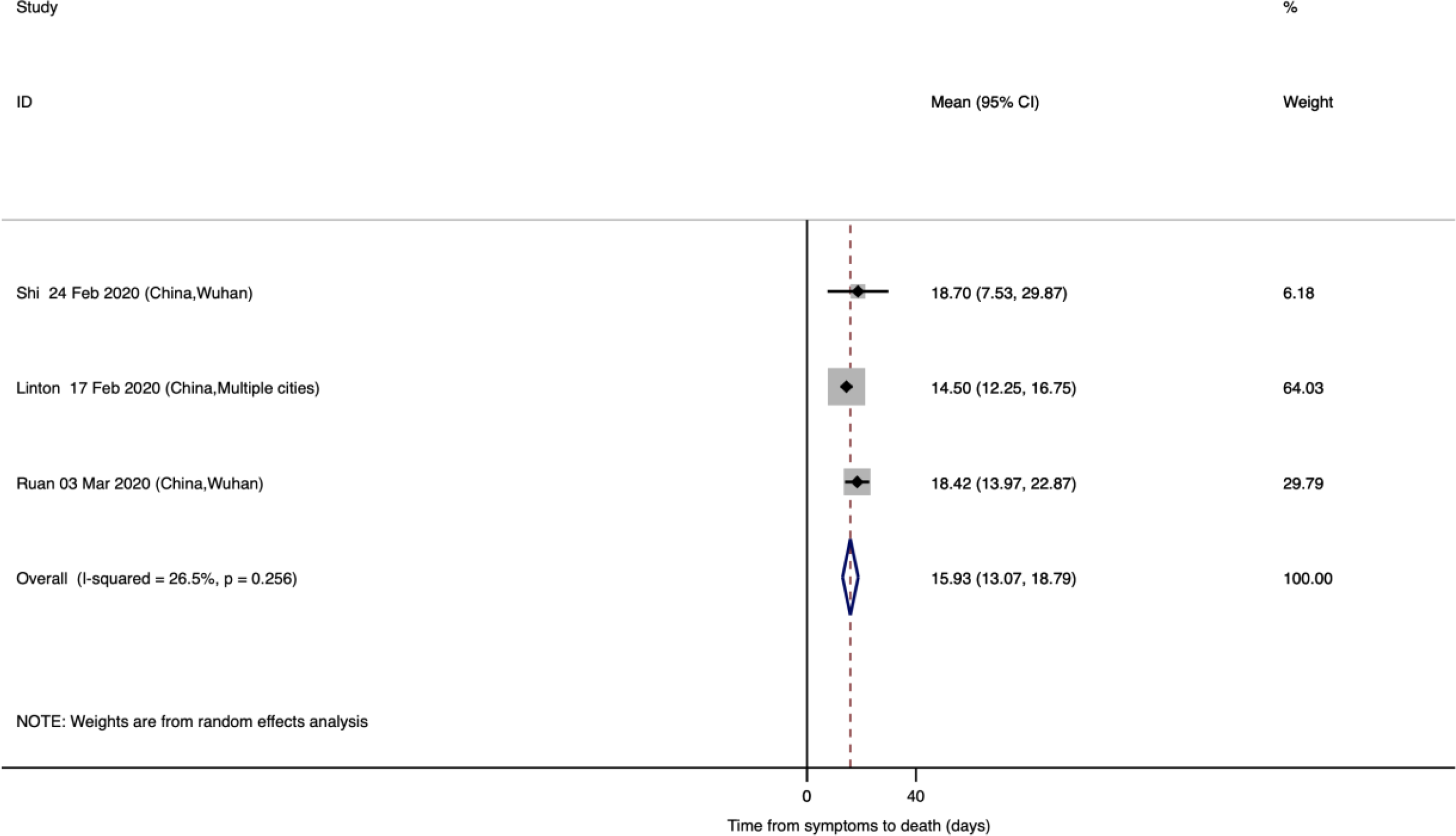
Time from onset of symptoms to death for COVID-19 infection

### Death probability

The estimated probability of COVID-19-related death was reported in 22 studies a pooled probability of 0.02 (95% CI: 0.02, 0.03) (Fig. 7). Heterogeneity testing (I^2^=97.6%) revealed notable differences among the included studies in the meta-analysis. Multivariate meta-regression results (**Supplement 2**) showed a significant difference in death probability by age (Adjusted Beta = 0.058; P-value = 0.016).

**Figure 7.**
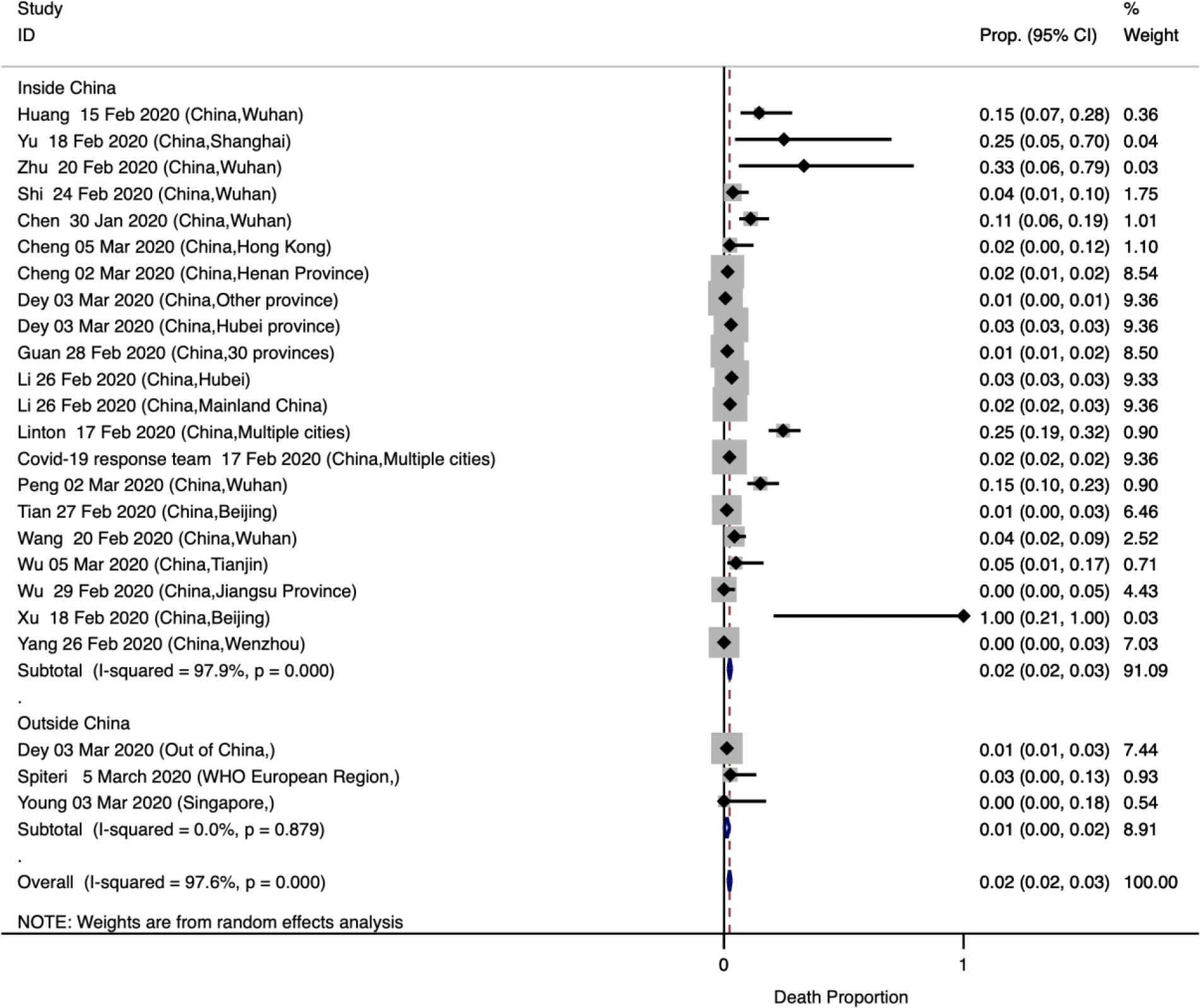
Probability of death among patients diagnosed with COVID-19 infection

## Discussion

We conducted a systematic review and meta-analysis to provide an overview of the epidemiological characteristics of COVID-19. Our findings suggested that the COVID-19 has an incubation time of more than five days (5.84 days) and a lag of less than five days (4.82 days) from onset of symptoms to first clinical visit. On average, the symptoms lasted more than twenty days (17.76 days) before recovery was achieved and the mortality risk among confirmed cases was 2%, which significantly increased by age. Similar to previous studies (21), fever, dry cough, shortness of breath, and fatigue were common symptoms among the patients in the included studies. As expected, history of direct or indirect exposure to Wuhan was frequent and the most common radiologic findings were bilateral consolidation and pneumonia (22, 23).

We found that the average incubation period of COVID-19 infection to be less than 6 days which is broadly consistent with previously reported estimates (24-27). The right tail of the incubation period for COVID-19 even for 99% CI was less than seven days (6.85). This finding is of particular interest as uncertainties continue to exist about the incubation period of COVID-19. For example, while both WHO and United States’ CDC suggest an incubation period of 2-14 days, single outlier cases as long as 19 (15), 24 (28) or 27 days (29) have been reported; estimates that are most likely reflecting a double exposure. Our meta-analytic findings are of particular importance for quarantine-related policies and planning and suggest that the current 14-day quarantine period might be rather conservative. Indeed, we found that except for one small study from China in Anyang city on a cluster of six patients (Bai 2020 (15)), all other studies reported incubation periods less than 11 days; therefore, a shorter period of 14 days would most likely suffice and it is expected that almost all infected people will show symptoms within 11 days of initial exposure. Nonetheless, decisions to modify or keep the existing policies need to weigh the costs of extending active quarantine against the potential or costs of missing a symptomatic case.

COVID-19 seems to have a longer incubation period than that of other acute respiratory viral infections (30) such as human coronavirus (3.2 days), influenza A (1.43-1.64 days (31)), parainfluenza (2.6 days), respiratory syncytial virus (4.4 days), and rhinovirus (1.4 days). Furthermore, the median incubation period for SARS has been reported estimated as 4.0 days in 2009 (30), which is considerably lower than what we observed for COVID-19. The longer incubation period of the COVID-19 may be one of the major factors that helps explain its rapid spread in comparison previous respiratory infection viruses. Another factor that contributes to spread of COVID-19 is the lag between the onset of symptoms and first clinical visit and the high number of asymptomatic cases of COVID-19. Our results indicated on average it is less than five days (4.82 days), and particularly increases by age (1.94 days increase per each 10 years increase in age). This finding suggests that MERS and SARS progress rapidly to sever symptoms and respiratory failures (32) than most cases infected by COVID-19 (33).

In compare to MERS-COV with a mortality rate of 35.67% (34) and SARS with a mortality rate of 11% (35), we found that COVID-19 has a much lower mortality rate (2%) which significantly increase by age (48% increase for every 1% increase in male participants). Although this estimate is comparable with previous studies (36, 37), it is important to recognize the limitations of calculating mortality rates of COVID-19 while the epidemic is still growing. As most cases of CVOID-19 remain asymptomatic and may recover without seeking medical care, it is likely that the true rate of death among people infected with COVID-19 could be even lower. On the other hand, the estimated mortality rates reported in most studies need to be interpreted with caution as they are often based on the cumulative number of deaths relative to the number of confirmed cases, while patients who die on a given day have been infected at a much earlier date and this would bias the denominator of the mortality rate (38).

We acknowledge four main limitations of our systematic review. First, our findings are mainly based on studies that recruited patient from clinics and hospitals and so may be biased toward more severe cases. Second, many studies did not report the study outcomes by subgroups such as age or gender and so we could report group-specific outcomes. Third, we used the mean and the standard error of the incubation period assuming a normal distribution which may have led to underestimate the right tail of the distribution. Lastly, given the urgency of topic and the heterogeneity of the studies included in the review, we did not conduct risk of bias and quality assessment of the studies. Inevitably, given the novelty of COVID-19 and the observational nature of all of the available evidence, they are most likely at a high risk of bias and the quality of existing evidence is low. Nonetheless, our systematic review of literature provides an update on epidemiological characteristics of COVID-19 which can be helpful for decision making on prevention measures as well as modeling and cost-analysis purposes.

## Conclusions

Our findings of the overall epidemiological characteristics of COVID-19 have important insight for healthcare systems’ prevention and planning efforts. The incubation period (i.e., <11 days in most studies) and the lag between the onset of symptoms and diagnosis (i.e., <5 days) are longer for COVID-19 compared to other respiratory viral infections including MERS and SARS.

Nonetheless, current policies of 14 days of mandatory quarantine for everyone, might be too conservative and longer quarantine periods might be more justified for extreme cases. As effective vaccination or treatment for COVID-19 are yet to be developed, practicing the fundamentals of public health and prevention efforts such as social distancing and personal hygiene are critical and need to be emphasized and enforced further to reduce the transmission risk of COVID-19 to vulnerable populations.

## Data Availability

All data is presented in the paper

## Notes

### Competing Interest Statement

The authors have declared no competing interest.

